# Community-led monitoring as a results-based strategy for improving rights-based HIV service delivery: A mixed-methods case study from Blantyre, Malawi

**DOI:** 10.64898/2026.08.26.26361390

**Authors:** Misheck Dickson Banda, Maalila Malambo

## Abstract

Despite Malawi’s progress toward the UNAIDS 95-95-95 targets, facility-level rights-based challenges in HIV services persist, including stigma, discrimination and limited community participation. Community-led monitoring (CLM) has been promoted as an accountability mechanism, yet independent, facility-level evidence from urban settings remains scarce. This convergent parallel mixed-methods study assessed CLM at Ndirande and Limbe health facilities in Blantyre using a client survey (n=250), key informant interviews (n=12), and focus group discussions (three groups, 15 participants), totalling 277 participants. Chi-square tests (with Cramér’s V) and binary logistic regression were used for the quantitative data; qualitative data were thematically analysed and triangulated. Analysis was guided by the rights-based approach to health and Arnstein’s ladder of citizen participation.

Awareness of CLM was moderate (56.0%) but participation was lower (40.2%), with involvement rated 2.78 out of 5, indicating consultative engagement. Awareness of CLM was the strongest and only robust predictor of participation (adjusted odds ratio ≈ 5.0, 95% confidence interval 2.4–10.6, p<0.001); a bivariate gender association did not survive adjustment. Notably, 41% of participants engaged in monitoring without recognising the term “CLM.” CLM strengthened community–provider communication (68.5%) more than responsiveness (36.2%). Accountability mechanisms existed but functioned informally and were inconsistently documented. The two facilities did not differ significantly on any of nine indicators (all p>0.12). Barriers were structural: funding, transport, staff attitudes, fear of reprisal, and cultural norms.

Urban CLM is a real but under-institutionalised accountability practice. The decisive lever is closing the awareness–action gap and formalising existing, unrecognised community monitoring through low-cost documentation, scheduled feedback, and independent, confidential complaint mechanisms. Findings are analytically transferable and offered as hypotheses for national piloting rather than as statistically generalisable conclusions.

## Introduction

HIV/AIDS is not only a health challenge but a development challenge, eroding household income, straining national budgets, and reducing the productivity of working-age adults, with effects that fall hardest on the poorest households and on women. The global response, articulated in the UNAIDS Global AIDS Strategy 2021–2026, has increasingly recognised that biomedical progress alone is insufficient, and that ending the inequalities which sustain the epidemic requires rights-based, people-centred and community-led approaches. Within this shift, community accountability mechanisms of which CLM is a leading example have moved from the margins to the mainstream of HIV programming, on the premise that affected communities are best placed to identify service failures and to press for their correction. Assessing whether this premise holds at the facility level, and under the demanding conditions of urban service delivery, is the task this study undertakes.

HIV/AIDS remains a major global public health challenge, with an estimated 40.8 million people living with HIV and 1.3 million new infections annually, and sub-Saharan Africa bearing approximately 65% of the burden [1,2]. Despite expanded antiretroviral therapy (ART) coverage, gaps in service quality, equity and accountability persist, particularly for marginalised and key populations [2,3]. The UNAIDS Global AIDS Strategy 2021–2026 emphasises rights-based, people-centred approaches that prioritise community leadership and accountability [4]. Malawi has achieved the UNAIDS 95-95-95 targets, with adult prevalence declining to 8.9% and annual new infections falling from around 56,000 in 2010 to approximately 12,000 in 2023 [5,6]. Yet national achievements mask persistent facility-level challenges in service quality and rights protection, which are acute in high-volume urban facilities such as Ndirande and Limbe in Blantyre, where the Southern Region records prevalence above the national average [6].

The evidence base on community accountability offers grounds for both optimism and caution. Systematic reviews of social-accountability interventions community scorecards, citizen report cards, community-based monitoring find that such mechanisms can improve service utilisation, provider effort and, in some cases, health outcomes, but that effects are highly heterogeneous and contingent on context: on the responsiveness of providers, the strength of feedback channels, the capacity and independence of community actors, and the wider institutional environment [7]. CLM inherits both the promise and the contingency of this broader family of interventions. Its distinctive emphasis on the leadership of affected communities aligns it with the long-standing principle in the HIV movement that those affected should be co-producers of accountability rather than passive beneficiaries, but whether that principle is realised in practice depends on the same enabling conditions that the general literature identifies. The present study contributes facility-level evidence on precisely these conditions in an urban African setting where they are least assured.

Community-led monitoring positions communities as active agents in monitoring and improving service quality, systematically collecting and analysing data on the availability, accessibility, acceptability and quality of services to advocate for improvements grounded in client experience [8,9]. Grounded in social accountability and the availability-accessibility-acceptability-quality (AAAQ) framework, and endorsed by PEPFAR, the Global Fund and WHO, CLM aims to bridge the gap between policy commitments and lived realities [10,11]. In Malawi, CLM has been implemented largely by civil-society organisations, and rural initiatives have reported positive outcomes such as increased multi-month ART dispensing [11]. However, most evaluations focus on rural settings, where traditional leadership and village structures facilitate mobilisation, and evidence on CLM’s facility-level functioning in urban areas, and on whether it genuinely shifts power between communities and providers, is limited [8]. This study addresses that gap by assessing CLM as a results-based strategy at two urban facilities. Its objectives were to assess community involvement in monitoring; evaluate rights-based service delivery; examine CLM’s influence on quality, accountability and responsiveness; and identify barriers. The study is grounded in the rights-based approach to health [12] and Arnstein’s ladder of citizen participation [13].

The rationale for focusing on urban facilities is both empirical and theoretical. Urban health centres in Malawi operate under conditions that differ markedly from rural ones: they serve larger and more mobile populations, experience higher patient volumes and longer waiting times, and lack the cohesive traditional structures chiefs, village heads, village development committees that facilitate community mobilisation in rural districts. The relative anonymity of urban life can weaken the social ties on which community accountability depends, while the heterogeneity of urban populations, including substantial numbers of key-population members, raises the stakes for equity and non-discrimination [11]. These features make urban facilities a demanding test of whether CLM can function where the conditions favouring community participation are least assured, and they justify treating the urban context as a distinct object of study rather than assuming that rurally-derived models will transfer.

Framing CLM as a results-based strategy is central to this study’s analytical approach. Results-based management orients activities toward measurable results through a continuous cycle of objective-setting, data collection, feedback, corrective action and review, and it places documentation at the heart of accountability. Evaluated through this lens, the pertinent question is not merely whether monitoring activities occur but whether they generate documented results whether community-generated evidence reliably feeds back into corrective action. This framing directs analytical attention to the integrity of documentation and feedback loops, which, as the findings show, is precisely where CLM at the two facilities is weakest.

## Materials and methods

### Design and setting

A convergent parallel mixed-methods design was used, guided by a pragmatist philosophy, at Ndirande and Limbe health facilities in Blantyre, Malawi’s commercial capital. Both are high-volume urban primary-care facilities serving densely populated catchments with HIV prevalence above the national average, and both host active CLM structures.

### Sampling

Purposive and stratified sampling were used. The minimum survey sample was estimated using Yamane’s formula (≈247); the achieved sample (250; Ndirande 128, Limbe 122) exceeded this minimum, and all valid responses were retained to maximise precision. Twelve key informant interviews (health staff, CLM focal persons, expert clients, a peer educator serving key populations, and a district coordinator) and three focus group discussions (15 community and Health Advisory Committee members) were conducted, giving 277 participants in total. Qualitative sample size was guided by data saturation.

### Data collection

Data were collected March–April 2026 using a KoboToolbox client survey, a key informant interview guide, and a focus group guide administered in Chichewa. The survey captured demographic characteristics, CLM awareness and participation, and perceptions of accessibility, equity, confidentiality, responsiveness and satisfaction.

### Statistical analysis

Quantitative data were analysed in RStudio (version 4.3). Associations between categorical variables were tested using the Pearson chi-square test with Cramér’s V as an effect size, and cells with low expected counts were flagged. Predictors of CLM participation were examined using binary logistic regression, reporting adjusted odds ratios with 95% confidence intervals; the model included gender, awareness of CLM, facility, age group and education as predictors, with reference categories as reported in Results. The significance threshold was set at α=0.05. A linear regression of satisfaction had been planned, but it was not pursued to inferential conclusions because the categorical predictor structure and the distribution of the satisfaction outcome did not meet the assumptions required for a stable model; satisfaction is therefore reported descriptively.

Qualitative data were transcribed, translated from Chichewa where necessary, and analysed thematically using a coding framework developed both deductively, from the study objectives, and inductively, from emerging themes. The two strands were integrated at the interpretation stage through triangulation, assessing convergence, complementarity and divergence in relation to each objective.

### Rigour and trustworthiness

Validity and reliability were pursued by deriving survey items directly from the study’s objectives and theoretical frameworks, administering the instrument electronically to reduce errors. Trustworthiness of the qualitative data was pursued through triangulation across three data sources and multiple informant types, the retention of original Chichewa quotations alongside translations, and an audit trail of coding decisions. Item-level missingness arising from skip logic was addressed transparently by reporting valid n per item rather than imputing values.

### Ethics statement

Ethical clearance for this study was obtained from the University of Malawi Research Ethics Committee (UNIMAREC), and permission to conduct the research was granted by the Blantyre District Health Office. All participants received clear information about the purpose of the study, their right to decline participation or withdraw at any time without consequence, and the confidential handling of their responses. All participants provided written informed consent before taking part. No minors were enrolled. Data were anonymised prior to analysis and are reported in aggregate.

## Results

### Socio-demographic characteristics

Survey respondents were predominantly female (62.0%) and aged 19–35 (54.4%); half (50.4%) were people living with HIV, and facility representation was balanced (Ndirande 51.2%, Limbe 48.8%) (Table 1). Most had at least secondary education (53.3%). Key informants (mean Community working experience 3.1 years) spanned both facilities and a range of roles, and focus group participants were aged 24–58.

**Table 1.** Socio-demographic characteristics of survey respondents.

| Characteristic | Category | n | % |
| --- | --- | --- | --- |
| Gender | Female | 155 | 62.0 |
|  | Male | 95 | 38.0 |
| Age | 19–35 | 136 | 54.4 |
|  | 36–60 | 66 | 26.4 |
|  | 15–18 | 36 | 14.4 |
|  | 61+ | 12 | 4.8 |
| Type | PLHIV | 126 | 50.4 |
|  | General user | 94 | 37.6 |
|  | Other/CLM member | 30 | 12.0 |

N = 250.

### Community involvement in monitoring

Awareness of CLM was moderate (135/241; 56.0%) but participation was lower (78/194; 40.2%), and involvement was rated 2.78/5 (SD 1.31; n=151), locating CLM at the consultative rung of Arnstein’s ladder. Awareness was propagated mainly through community meetings and non-governmental organisations rather than the facility (Table 2). Among participants, involvement was predominantly consultative attending meetings and advocacy actions rather than participation in decision-making review sessions (Fig 1).

**Fig 1.**
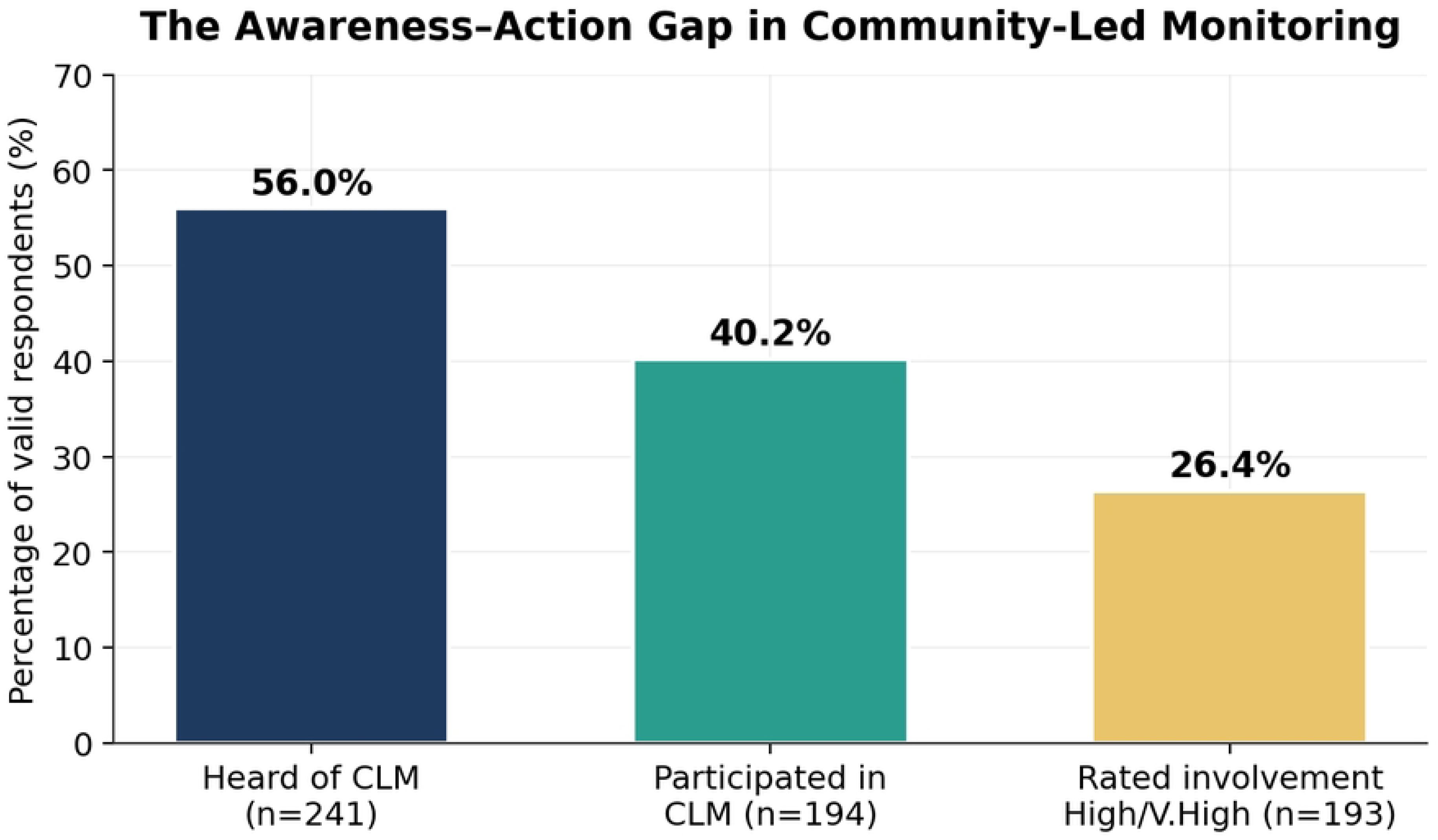
The awareness–action gap in community-led monitoring.

**Table 2.** Sources through which respondents first heard of CLM.

| Source of first awareness | n | % of aware (135) |
| --- | --- | --- |
| Community meeting | 58 | 43.0 |
| Non-governmental organisation | 44 | 32.6 |
| Health facility | 24 | 17.8 |
| Friend or relative | 24 | 17.8 |
| Radio, church or youth club | 14 | 10.4 |

Disaggregating the forms of involvement revealed the consultative character of participation. Among participants, the most common activities were attending community meetings (42.0%) and participating in advocacy actions (35.8%), followed by monitoring service quality (27.2%) and collecting feedback from users (21.0%); participation in review sessions with facility staff, the form most closely tied to decision-making, was least common (13.6%). Participants were mobilised chiefly by community leaders (32.1%), health workers (26.9%) and peer educators (23.1%).

Awareness of CLM was the strongest predictor of participation at the bivariate level (χ²=20.44, p<0.001, Cramér’s V=0.333); gender showed only a weak association (χ²=4.20, p=0.040, V=0.148) and facility none (p=0.975). In logistic regression (n=169), awareness remained strongly predictive (adjusted odds ratio [OR]≈5.0, 95% confidence interval [CI] 2.4–10.6, p<0.001), while gender did not survive adjustment (OR 1.65, 95% CI 0.82–3.32, p=0.16), indicating that the apparent gender effect largely reflects differences in awareness (Table 3). A novel finding was that 41% (32/69) of participants engaged in monitoring despite not recognising the term “CLM,” indicating that community monitoring practice runs ahead of formal CLM literacy (Fig 2).

**Fig 2.**
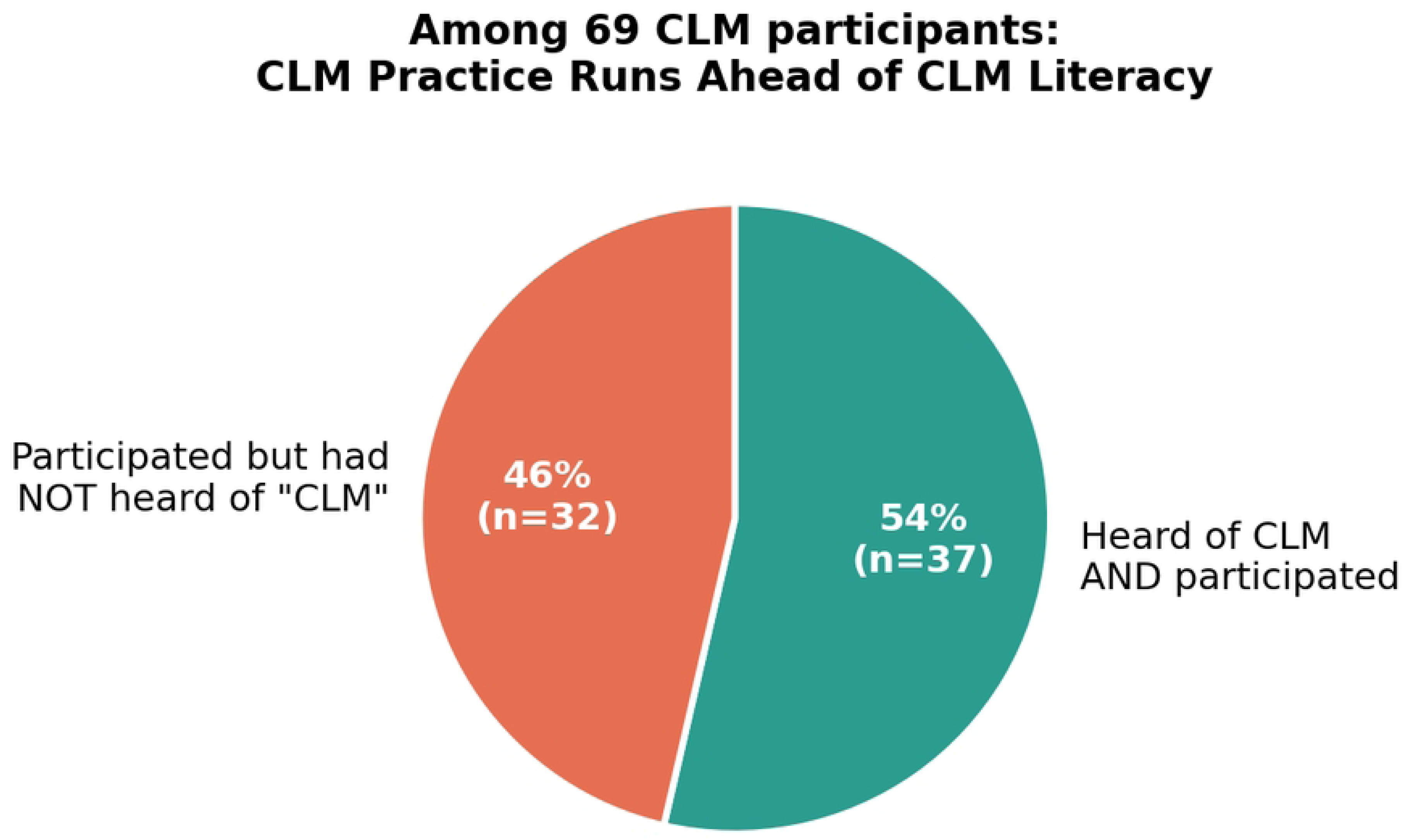
CLM practice ahead of CLM literacy.

**Table 3.** Binary logistic regression predicting CLM participation.

| Predictor | Adjusted OR | 95% CI | p |
| --- | --- | --- | --- |
| Heard of CLM (ref: No) | 5.02 | 2.38–10.6 | <0.001 |
| Female (ref: Male) | 1.65 | 0.82–3.32 | 0.16 |
| Ndirande (ref: Limbe) | 0.81 | 0.40–1.62 | 0.55 |
| Age 36–60 (ref: 19–35) | 1.29 | 0.57–2.91 | 0.55 |

$n=169$ ; McFadden pseudo- $R^2=0.10$ .

### Rights-based service delivery

Access was rated easy or very easy by 60.6% (of 241), with the main barriers being lack of privacy, transport costs, unfriendly staff, long waiting times and stigma. Equity was the weakest dimension: only 38.4% felt clients are always treated equally, and 31.2% had witnessed discrimination, most commonly on grounds of disability, age, gender and HIV status, and disproportionately affecting key populations (Fig 3). Among those answering confidentiality items (n=126), 37.3% felt information was always confidential, but 22.2% had avoided services for fear of breaches. Overall rights-based satisfaction was moderate (mean 3.42/5, SD 1.22). The right to remedy was compromised by fear of reprisal, indicating that low complaint rates reflect under-reporting of grievances.

**Fig 3.**
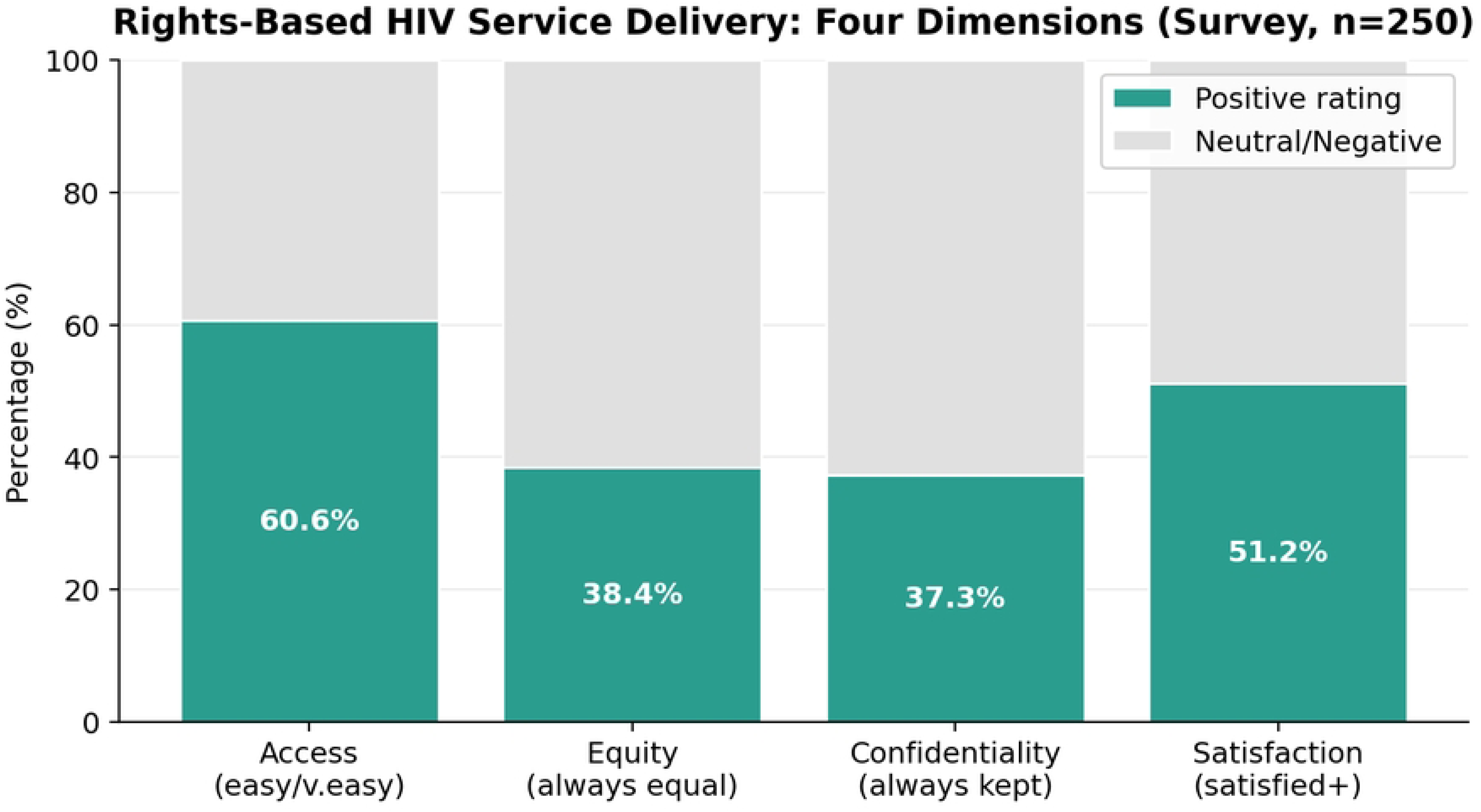
Rights-based service delivery across four dimensions.

### Influence of CLM

Perceived improvements reduced waiting times, coded registration replacing open registration, renovated youth-friendly spaces, and Saturday adolescent clinics were reported consistently across independent key informants but were concentrated among those directly involved in CLM. Among respondents answering the relevant items, 68.5% (of 54) felt CLM improved communication, while only 36.2% (of 116) felt it improved responsiveness, and satisfaction with facility response was moderate (mean 2.95/5). Accountability mechanisms existed (47.8% had seen them) but functioned informally: suggestion boxes were often unopened, meetings irregular, and written records inconsistently maintained across sites a documentation weakness rather than a total absence. Where feedback was documented and acted upon, the results were tangible; where it was not, issues recurred.

### Barriers and facility comparison

Barriers, synthesised from the qualitative sources, were inadequate and delayed funding, transport costs for monitors, staff attitudes and turnover, fear of stigma and reprisal, cultural and gender norms, and a shortage of documentation materials (Fig 4). These barriers were mutually reinforcing, forming a cycle in which resource scarcity weakened documentation, weak documentation undermined accountability, and invisible accountability eroded community trust. The two facilities did not differ significantly on any of nine indicators (all p>0.12), indicating systemic rather than site-specific constraints (Fig 5) (Table 4).

**Fig 4.**
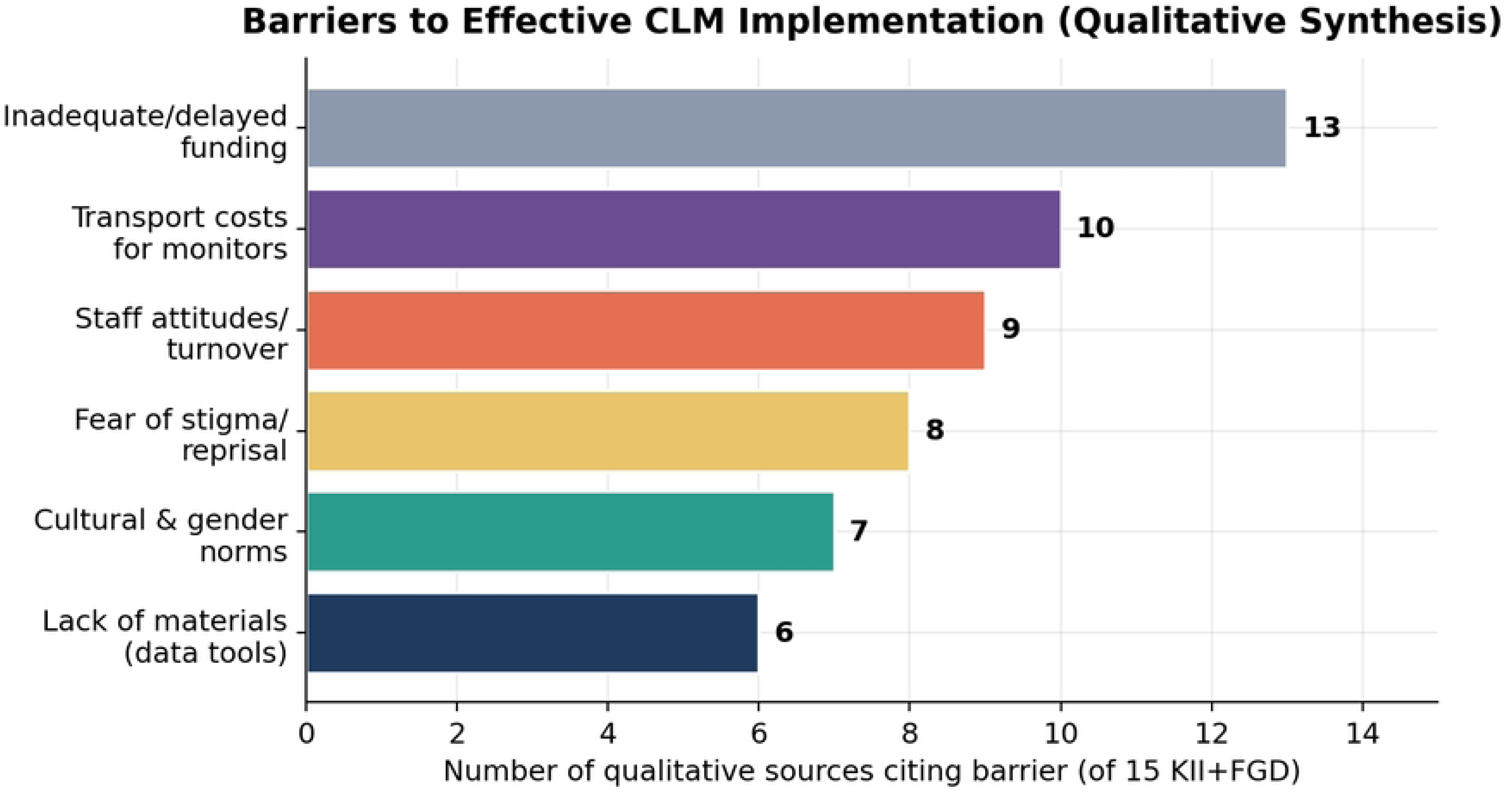
Barriers to CLM implementation.

**Fig 5.**
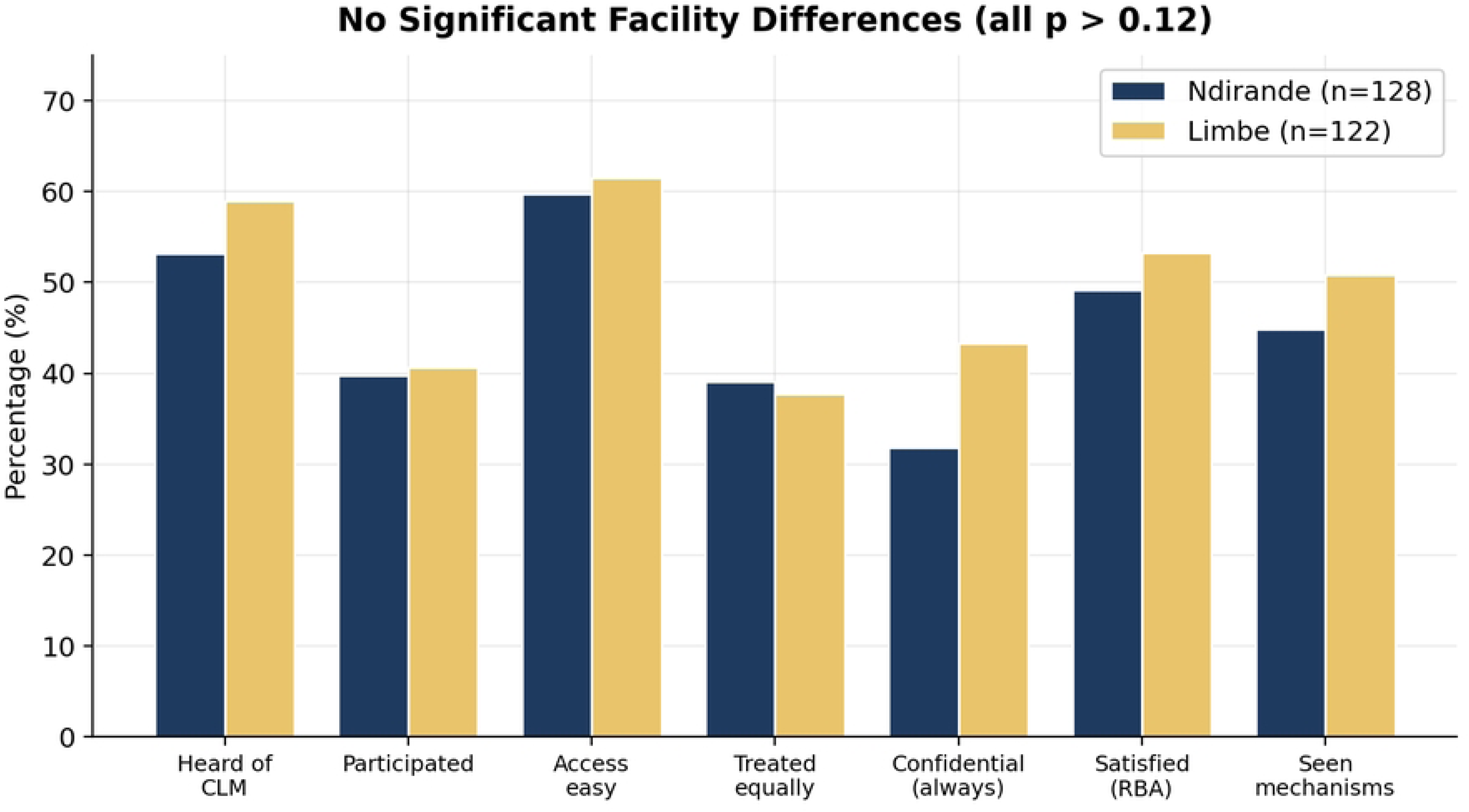
Facility comparison across nine indicators.

**Table 4.** Facility comparison (chi-square). No indicator was statistically significant.

| Indicator | Ndirande % | Limbe % | p |
| --- | --- | --- | --- |
| Heard of CLM | 53.2 | 59.0 | 0.44 |
| Participated | 39.8 | 40.6 | 1.00 |
| Access easy | 59.7 | 61.5 | 0.87 |
| Always treated equally | 39.1 | 37.7 | 0.93 |
| Confidential (always) | 31.8 | 43.3 | 0.25 |
| Satisfied (rights-based approach) | 49.2 | 53.3 | 0.61 |
| Seen mechanisms | 44.9 | 50.8 | 0.42 |

## Discussion

This study provides one of the first independent, facility-level assessments of CLM in urban Malawi. Its central contribution is to reframe the participation deficit. With awareness the decisive predictor of participation, and 41% of participants unaware of the CLM label, the binding constraint is less community apathy than the gap between widespread but unrecognised monitoring activity and the awareness, resourcing and documentation needed to formalise it. The highest-yield intervention is therefore to name, label and institutionalise the community activity that already exists rather than to build participation from nothing. This reframing has, to our knowledge, not been documented in the Malawian literature, and it carries direct implications for programme design, since studies that measure participation only among those recruited into formal CLM structures will systematically underestimate community monitoring and miss the population for whom formalisation, rather than mobilisation, is the appropriate response.

Consistent with evidence from South Africa and Uganda [11], CLM was associated with tangible improvements in waiting times, commodity availability and staff attitudes. The consistency of these accounts across independent informants at both facilities strengthens their credibility. However, CLM’s stronger effect on communication than on responsiveness indicates that it opens channels of voice faster than it closes feedback loops into documented action. Interpreted through results-based management, the cycle breaks at the point of documentation: CLM strengthens data collection and feedback but not the recording, action-tracking and review that convert voice into durable results. This diagnosis is more precise than a binary claim that documentation is absent, and it points to a correspondingly precise, low-cost remedy.

The consultative depth of participation locates CLM within the tokenism band of Arnstein’s ladder, consistent with the social-accountability literature’s caution that participatory mechanisms often leave the underlying distribution of power untouched [7]. Yet the coexistence of consultative participation with real service improvements suggests that even tokenistic participation can yield gains where facilities are receptive, supporting a developmental rather than a dismissive view of CLM. The equity findings persistent discrimination and fear-driven under-use indicate that the technical successes of the HIV response have outpaced progress on the interpersonal and rights dimensions of care, underscoring the continued relevance of the rights-based approach. The finding that fear of reprisal suppresses complaints reveals that the right to remedy fails not for want of complaint channels but for want of trusted, independent ones.

The statistical equivalence of the two facilities across all nine indicators is analytically pivotal: it indicates that the constraints on CLM are systemic to the urban primary-care context rather than the product of local management, and it therefore directs interventions to the district and national levels. This equivalence also enhances the transferability of the findings to other urban facilities facing the same systemic conditions, while cautioning against constructing a “better facility” narrative unsupported by the data.

The confidentiality findings merit particular emphasis because of their behavioural consequences. That 22.2% of respondents had avoided seeking services at some point for fear of confidentiality breaches represents a direct threat to the HIV response: avoidance undermines treatment continuity, retention in care and, ultimately, viral suppression. The qualitative evidence that CLM had driven a shift from open to coded registration and the creation of private consultation spaces demonstrates that community monitoring can address such concerns concretely, yet the persistence of recalled breaches shows that the underlying risk has been reduced rather than eliminated.

The barriers identified funding, transport, staff attitudes and turnover, fear of reprisal, and cultural and gender norms correspond closely to those documented across the sub-Saharan literature on community health worker and social-accountability programmes, suggesting that they are structural features of community monitoring in low-resource settings rather than local peculiarities. This study’s contribution is to show how these barriers interlock into a self-reinforcing cycle centred on the documentation deficit: without funding and materials, documentation weakens; without documentation, accountability remains informal and person-dependent; and without visible accountability, the community trust on which participation depends erodes.

The divergence between the survey and qualitative findings on the depth of monitoring activity is analytically productive rather than contradictory. The survey, which measures recognition of the formal CLM label, portrays a shallow reach, while the qualitative data, which captures lived practice, portrays a richer set of monitoring activities already under way. Reconciled, the two strands yield the study’s central insight: communities are conducting more monitoring than they recognise as “CLM,” and the task of policy is to name, formalise, resource and document that activity rather than to create participation from nothing.

### Implications

Four priorities follow: (1) close the awareness–action gap through deliberate, facility-based CLM communication and by connecting with the informal networks through which communities already encounter CLM; (2) institute simple, low-cost documentation complaint registers, meeting minutes, action trackers to close the results-based cycle; (3) establish independent, confidential complaint mechanisms, and actively promote awareness of them, to counter fear of reprisal; and (4) secure sustainable funding by embedding CLM in district and facility budgets and providing for monitors’ transport and materials.

### Limitations

The study was confined to two urban facilities, limiting statistical generalisability, particularly to rural settings which was the deliberate design of the study. The cross-sectional design precludes causal inference; purposive sampling and self-report introduce potential bias; and documentary evidence was incomplete, so accountability findings rely on triangulated participant accounts. Because the survey measured recognition of the CLM label while interviews captured lived practice, part of the practice–literacy gap could reflect an instrument effect, though convergent evidence supports the substantive interpretation. Findings are therefore offered as analytically transferable propositions for national piloting.

## Conclusions

CLM at Ndirande and Limbe represents a real but incomplete shift toward community accountability in HIV service delivery. It has strengthened communication and contributed to measurable service improvements, but consultative participation, informal and inconsistent documentation, and fear of reprisal constrain its transformative potential, holding it at the consultative rung of Arnstein’s ladder. Realising its potential requires formalising the community monitoring that already exists through awareness-raising, reliable low-cost documentation, independent complaint mechanisms, and sustained funding to advance CLM from consultation toward genuine citizen power. These findings offer a credible, evidence-based agenda for strengthening community-led accountability in urban HIV services in Malawi and comparable settings.

## Data Availability

The minimal de-identified dataset underlying this study's findings is available at Zenodo via https://doi.org/10.5281/zenodo.21562464

## Acknowledgments

The authors thank the management and staff of Ndirande and Limbe health facilities, the Blantyre District Health Office, and all survey, interview and focus group participants for their time and openness in sharing their experiences of community-led monitoring and HIV service delivery.

